# Characteristics and Impact of Librarian Co-authored Systematic Reviews: A Bibliometric Analysis

**DOI:** 10.1101/2020.02.14.20023119

**Authors:** Ahlam A. Saleh, Frank Huebner

**Affiliations:** Health Sciences Library, University of Arizona, Tucson, Arizona, United States; Independent Researcher, Tucson, Arizona, United States

**Keywords:** Librarians, Information specialists, Libraries, Systematic reviews, Meta-analyses, Bibliometrics, Impact, Authorship

## Abstract

**Background:** Health sciences libraries have been providing services that support systematic reviews (SRs) for many years. In recent times the problem facing health sciences libraries is the management of the demand versus resources availability. There have been questions posed as to the value of this type of service in health sciences libraries. A valuable outcome of librarian collaboration on SR teams is co-authorship of the reported SRs. This study aimed to examine the characteristics and impact of librarian co-authored SRs.

**Methods:** A bibliometric analysis was conducted. Librarian co-authored SRs were identified in the Web of Science (WOS) Core Collection limited up to the year 2017. Librarian co-authored SRs with the librarian as first author were excluded from this analysis. Additional inclusion and exclusion criteria were applied in the selection process. The included records were analyzed using Perl programs and VOSviewer. To examine the dissemination of librarian co-authored SRs, citing articles to the included SR records were retrieved from the WOS Core Collection and then identified in MEDLINE for an analysis of the indexed publication types.

**Results:** The included results yielded 1,711 librarian co-authored SRs published between 1996 and 2017. The top three countries of the first author affiliation were USA, Canada, and Netherlands. Sources of publication were distributed among 730 journal titles. The number of MEDLINE citing articles to the included SRs was 28,868. The mean number of citations to a SR was 26.4. The top publication type descriptor of the citing articles representing the MEDLINE “Study Characteristic” category was “Randomized Controlled Trial”.

**Conclusion:** Outcomes of librarian contributions to supporting SRs include increasing scholarship opportunities that highlight librarian contributions to other disciplines. SRs are bodies of evidence, which can influence policy, patient care, and future research. In this study, we demonstrate that librarian co-authored SRs are disseminated into randomized controlled trials and other study types, meta-analyses, as well as guidelines, thus providing insight into knowledge transfer and the potential for clinical implementation.

## INTRODUCTION

With their expertise in searching and information management, librarians are well positioned to support the production of systematic reviews (SRs). This expertise is recognized by external stakeholders as is evident by the recommendation to include librarians and information specialists in the SR process, as outlined in the Institute of Medicine (IOM) standards on conducting SRs, *Finding What Works in Health Care*, released in 2011 [1]. Many librarians across different professional settings are engaged in SRs, and supporting SRs has become increasingly standard in the scope of work performed by health sciences librarians. The growing role of librarians in SRs is reflected in the extensive programming on this topic at Medical Library Association annual meetings and continuing education efforts and is the subject of a recent scoping review [2].

SRs are labor- and time-intensive. When SR support is just one of many other job responsibilities, as is the case in most academic and hospital settings, librarians and libraries can become overwhelmed with the demand for SR services, and struggle to find the best way to balance workloads. The increasing demand in requests for SR support has led some library administrators to question the value of providing such a service [3], and some libraries have even implemented a pay-per-service model [4, 5].

Another source of SR-related stress is the fact that in some academic libraries with tenure tracks, the contributions of librarians who successfully collaborate on SRs as co-authors, including coordinating large sections of the SR process, writing portions of the manuscript, and editing manuscripts, may not be considered to meet scholarship requirements because they do not directly focus on a topic within the field of library and information science (LIS). Thus, further research exploring librarians’ substantive role in SRs would give the information professional community a better understanding of the scholarly contributions and impact of this type of work.

Previous research in the LIS literature examined the added value of librarians’ contributions to the quality of SRs. Meert et al. identified SRs in which librarians contributed as co-authors or team members and scored those SRs using a 15-item reporting methodology checklist. They found that librarian involvement correlated with a higher quality score for the literature search component of SRs [6]. Rethlefsen et al. found that librarian/information specialist co-authored SRs reported higher-quality search strategies [7]. Also, Koffel found a positive correlation between librarian involvement in SRs and the use of SR guideline-recommended search methods [8]. These studies provide evidence of the value of librarian contributions to performing and reporting SR searches. However, there is limited published research on other characteristics and the overall impact of librarian co-authored SRs.

Bibliometrics is “the application of mathematics and statistical methods to books and other media of communication” as coined by Pritchard in 1969 [9]. Data regarding research output, impact, and collaborations are some examples of bibliometric measures that can be valuable in strategic decision-making and research assessment. While citation analysis alone provides insight into research impact, combining citation analysis with qualitative methods and analyses can provide greater context and improve its usefulness. Research assessment models such as the Becker Model [10] were developed to provide a framework for highlighting the different ways by which research impact can be captured.

Citation analysis as a method to assess impact has been used in previous LIS literature investigating the value of medical library services on healthcare literature. For example, Sherwill-Navarro et al. conducted a citation analysis and examination of the dissemination of four selected library services research studies to demonstrate impact [11]. However, this study did not look at SRs specifically nor aimed to achieve evaluation of the scope of studies analyzed that our study aimed to do. Regarding bibliometrics in the healthcare literature, Royle et al. conducted a bibliometric analysis of SRs and further analysis to determine potential predictors of the citation impact; and Rosas et al. used bibliometric analysis and network visualization to evaluate the research and impact of National Institute of Allergy and Infectious Diseases (NIAID) publications [12, 13]. Neither of these studies looked at librarian co-authored SRs, specifically. Furthermore, Rosas et al. examined the uptake of the NIAID publications in research reviews and syntheses [13]. In our research, we also went beyond citation analysis counts to examine what types of publications the SR research we studied were cited in and similarly we conducted visual mapping analysis as part of the bibliometric methods. Given the limited published research on the scholarly contributions and impact of librarians’ work related to their collaborative role in the production of SRs, we sought to demonstrate the impact of librarian co-authored SRs and to identify characteristics of librarian co-authored SRs. In this study, we went beyond conducting citation analysis to examine where research was cited to shed light on the diffusion of SR research. This type of content analysis along with citation analysis provides us with a more informed view about the impact of librarian co-authored SRs.

## METHODS

### Retrieval and selection of SR records

For the purposes of this study, we defined “librarian” as a co-author with a library affiliation as identified by the address field in the Clarivate Analytics Web of Science (WOS) record, our designated source for identifying the SRs in this study.

Inclusion criteria for our study were as follows: (1) the record indicated that a SR or meta-analysis was conducted or the source of publication was the Cochrane Database of Systematic Reviews (full text was consulted if any clarification was needed), and (2) a librarian was a co-author as determined by affiliation with a library (*lib* or *library* or *libraries* in the address or organization field in the WOS Core Collection record; full text and/or websites were consulted as needed for clarification). Exclusion criteria were as follows: (1) the publication was a letter, comment, reply, or opinion piece; (2) the publication was a protocol for a SR or meta-analysis; (3) a librarian was the first author; (4) the affiliation was unclear; (5) it was a duplicate citation.

Works for which the librarian was first author were not included in this review because we aimed to specifically examine the role of librarian as collaborator providing their expertise to support non-LIS discipline SRs.

The WOS Core Collection was used to retrieve SR records in October 2018. The following subsets of the WOS Core Collection were used: Science Citation Index Expanded (1900-present), Social Sciences Citation Index (1900-present), Arts & Humanities Citation Index (1975-present), Conference Proceedings Citation Index-Science (1990-present), and Conference Proceedings Citation Index-Social Science & Humanities (1990-present). The search strategy used to retrieve SRs was: (ORGANIZATION-ENHANCED: (lib OR library OR libraries) OR ADDRESS: (lib OR library OR libraries)) AND (TITLE: (“systematic review” OR “systematic literature review” OR “meta-analysis” OR metaanalysis) OR PUBLICATION NAME: (Cochrane Database of Systematic Reviews)). To gauge trends over time, searches included records from the inception of the database, as indicated above. To allow for a reasonable time frame for the literature to accrue citations, search results were limited to those published in or before 2017. Search results were also limited to the following document types: Review OR Proceedings Paper OR Article OR Meeting Abstract. We included conference material such as proceedings papers and meeting abstracts as these are also examples of scholarly contributions. Document types that were excluded were: Early Access (“articles in press,” “published ahead of print”), Editorial Material, Letter, Book Chapter, and Book.

Prior to conducting the screening process, we piloted the inclusion/exclusion criteria and made modifications when we felt further clarification was needed. Search results from WOS were exported into EndNote (version X8.2). One author screened the records based on our inclusion and exclusion criteria.

### Analysis of included SRs

Bibliometric mapping allows visualization and linking of bibliometric data, which can aid in illustrating geographic, network, and prominent terms that provide insight into the research areas represented by the records. We used the VOSviewer program (versions 1.6.10 and 1.6.11) to analyze the research areas and citation impact of the selected SRs [14]. For research area analysis, a map was created based on co-occurrence of terms from the title and abstract fields of the records. We used the binary counting setting and designated a threshold of a minimum of 15 appearances across records and 60% relevance (i.e., to eliminate irrelevant, general terms that do not provide useful information) for a term to appear in the map. Further information on relevance determination using VOSviewer is described by Van Eck and Waltman [15]. Additional irrelevant terms and synonyms were rectified using a thesaurus file for VOSviewer.

### Analysis of citing article records

The included SR records were then located in WOS, and a citation report was created to identify their citing articles. The citing articles (including self-citations) were exported in full record plain text file format. The inclusion of self-citations is a matter of ongoing debate in citation analysis. However, we included self-citations because they are a natural part of scholarly communication; researchers build on their previous work and should not necessarily be assumed to intentionally inflate citations of their own publications [16, 17]. Because of the large number of records (>41K), we utilized Perl programs to perform data analyses.

As a result of the large scale of data requiring analysis, we used a set of authority terms indexed to records instead of manually characterizing the records ourselves. Specifically, we used the National Library of Medicine Medical Subject Heading (MeSH) descriptors for Publication Characteristics, as they provide a much richer description of publication types compared to those provided by WOS. Throughout the remainder of the manuscript, we use the term “publication type” to refer to these descriptors, which are indexed in the Publication Type data element (field) available in the MEDLINE subset of records in PubMed.

We classified the publication types of citing articles according to MeSH categories falling under the Publication Characteristics (Category V Descriptors) section of the MeSH tree hierarchy [18]. The descriptors listed under Publication Characteristics are organized into the following four categories: “Publication Components”, “Publication Formats”, “Study Characteristics” and “Support of Research”.

PubMed unique identifiers (PMIDs) were obtained from the citing article records using a Perl program and then entered into PubMed to retrieve the citing articles available in PubMed. If there was no PMID in the WOS citing article record, the record was excluded from further analysis. PMID search results in PubMed were then limited to MEDLINE records only, with non-MEDLINE records being excluded from further analysis. Results were limited to the MEDLINE subset of records because non-MEDLINE records may have incomplete or inaccurate publication types [19]. The resulting MEDLINE records were exported and input into a Perl program to perform analysis.

## RESULTS

The WOS search for librarian co-authored SRs yielded an initial 1,817 records (Figure 1). Of these, 1,711 records were selected as SRs co-authored by librarians according to our inclusion and exclusion criteria.

**Figure 1.**
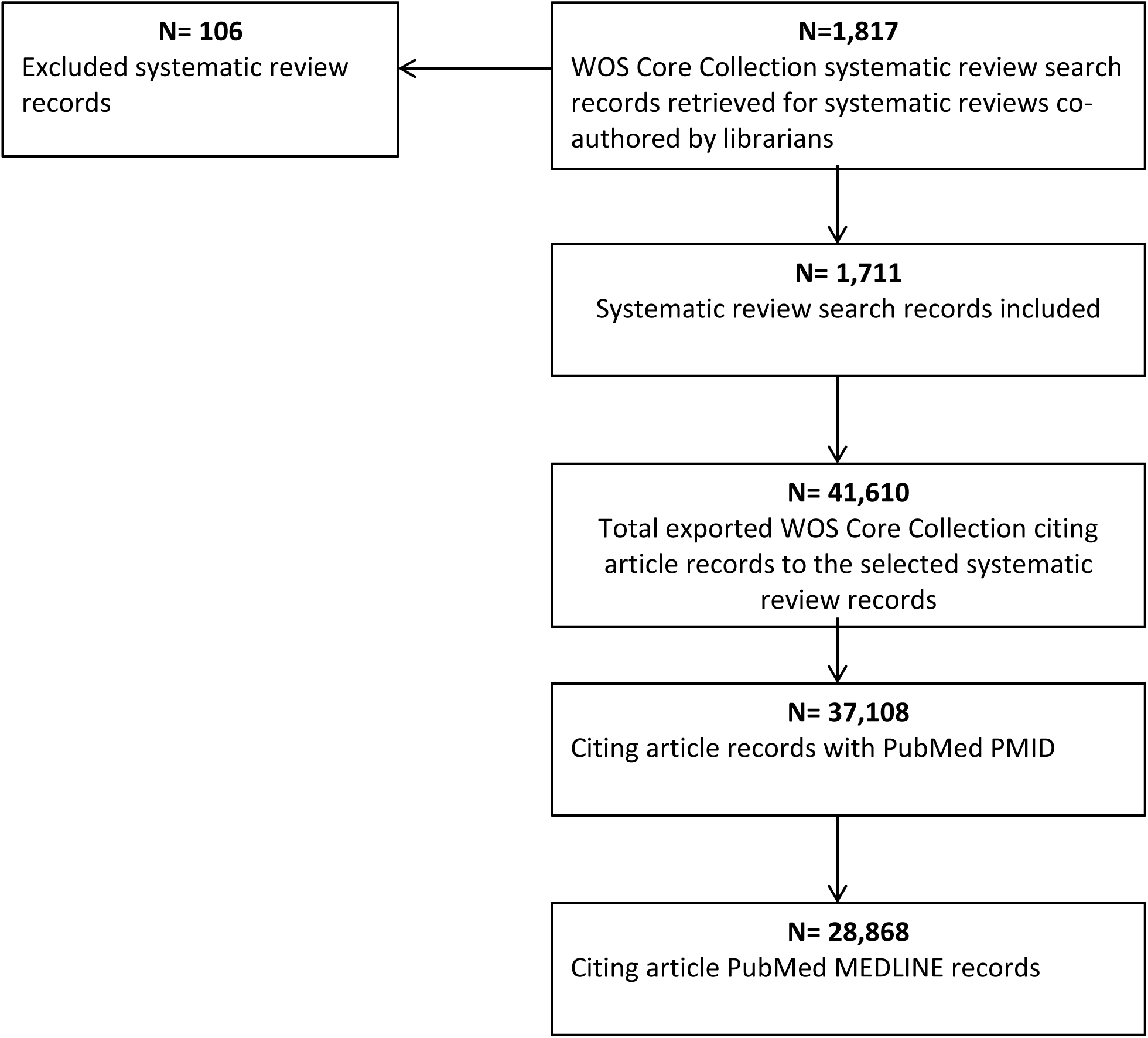
Flow chart of the retrieval and selection of records.

### Characteristics of librarian co-authored SRs

In VOS Viewer, of the 33,100 terms identified in the 1,711 SRs, 350 met the frequency threshold, and 210 were sufficiently relevant to be selected for visualization. The density visualization map in Figure 2 depicts the topic areas of the SRs. The most common types of SRs appeared to be for interventions and diagnostic test accuracy, and the two most represented disease categories were “cancer” and “cardiovascular disease”. The overlay visualization map in Figure 3 depicts the normalized citation impact of the SRs. Normalized citation impact corrects for the age of a document defined by VOSviewer as follows: the “normalized number of citations of a document equals the number of citations of the document divided by the average number of citations of all documents published in the same year and included in the data that is provided to VOSviewer” [20]. Our overlay visualization map illustrating citation impact shows that SR topics such as cardiovascular disease, obesity, diabetes, and training are particularly impactful.

**Figure 2.**
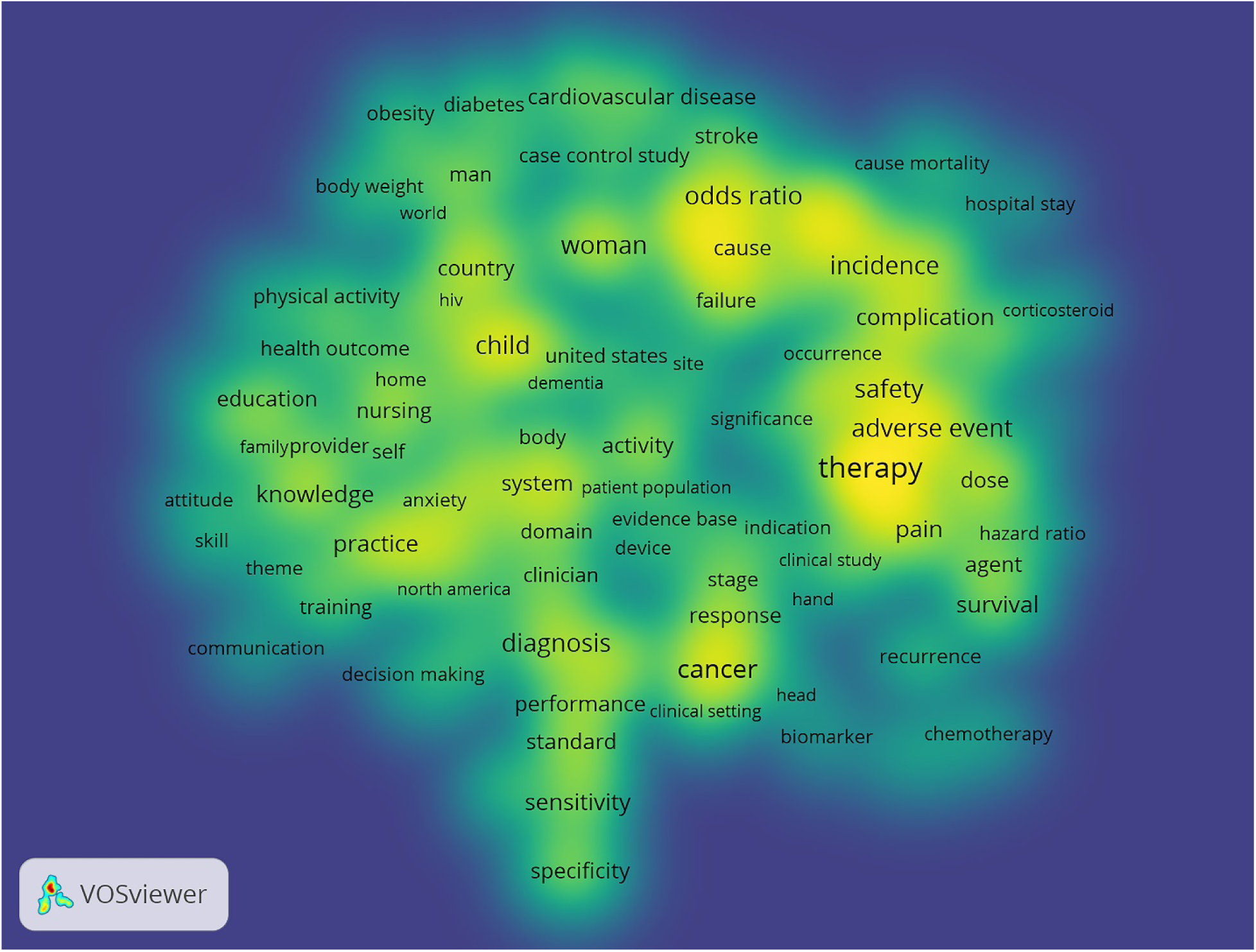
Density visualization map of librarian co-authored SRs. Term coloring is proportional to the number of term occurrences, with yellow representing the highest frequency of occurrences and blue representing lower frequencies. Font size is also proportional to the number of term occurrences, with larger sizes indicating higher frequencies.

**Figure 3.**
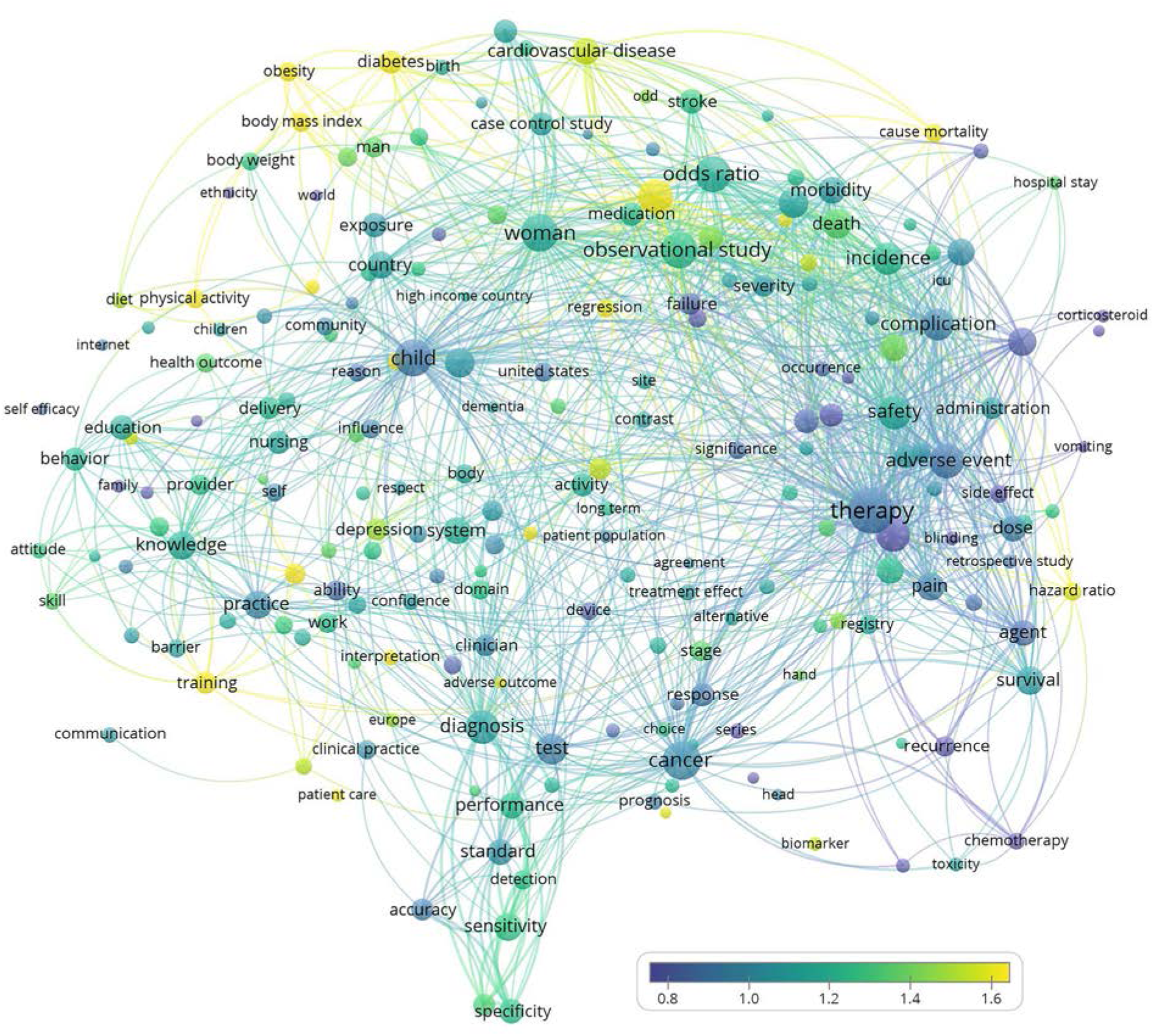
Overlay visualization map of librarian co-authored SRs. Circle size reflects the number of term occurrences across records. Lines between circles indicate term co-occurrence across records. Circle color reflects the average normalized citation impact of all records in which the term occurs in the title or abstract. Blue indicates lower citation impact, and yellow indicates higher citation impact.

The number of times librarian co-authored SRs were cited ranged from 0 to 1,012, with an average of 26 and a median of 10. Of the 1,711 SRs, 187 (10.9%) were uncited, which comprised of 65 (3.8%) of the WOS document types “article” or “review”; 120 (7.0%) of the WOS document type “meeting abstract”; and 2 (0.1%) of the WOS document type “proceedings paper”. Also, 41.7% of the uncited SRs were published in 2017. The number of authors on the SRs ranged from 2 to 39, with an average of 7 authors. Additional characteristics of the SRs are outlined in Table 1.

**Table 1.**
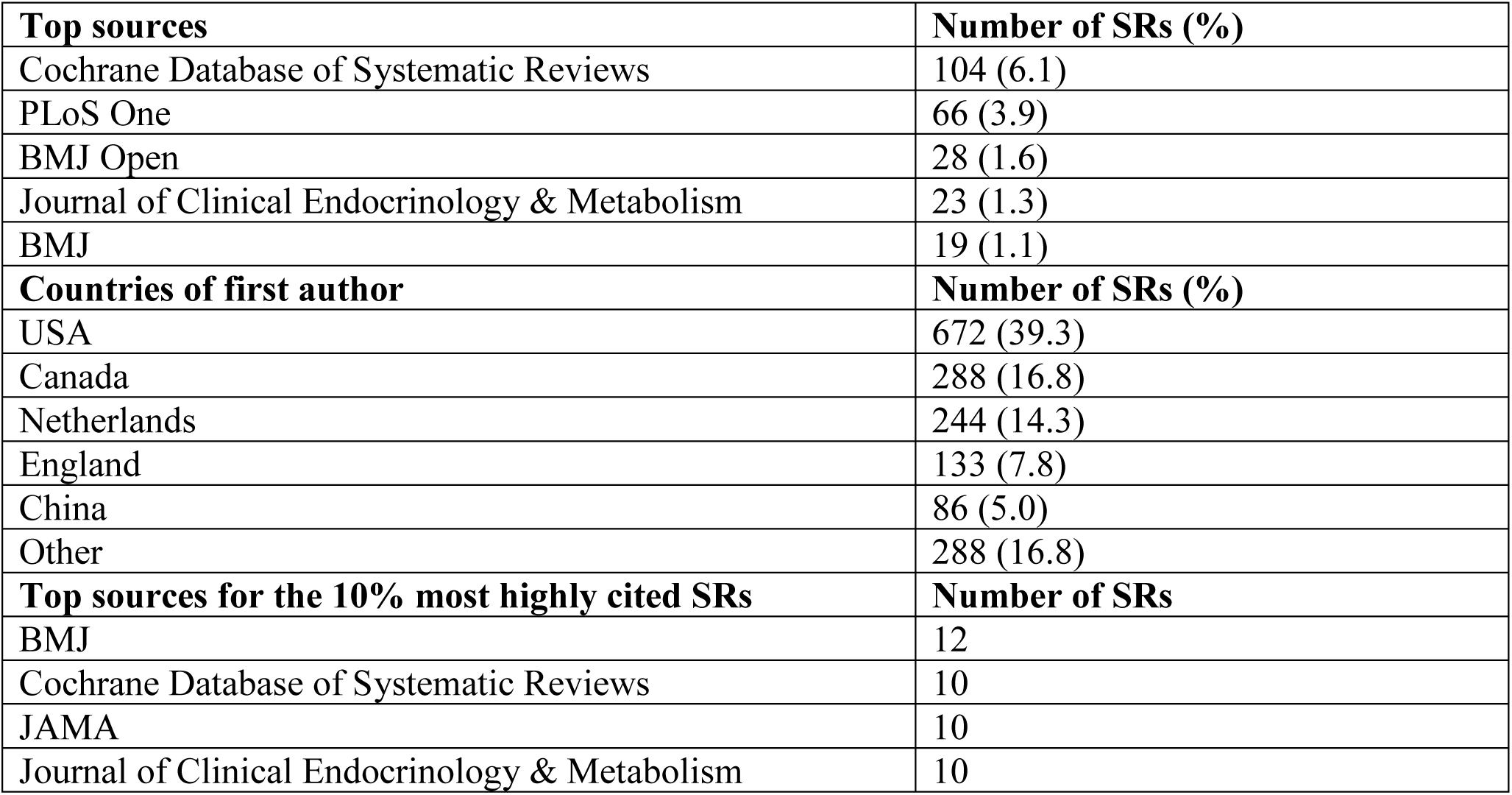
Characteristics of the librarian co-authored SRs.

The number of SRs published over time from 1996-2017 is shown in Figure 4. The temporal trend demonstrates the start of an increasing number of librarian co-authored SRs in the year 2006 with a steeper increase beginning in the year 2011.

**Figure 4.**
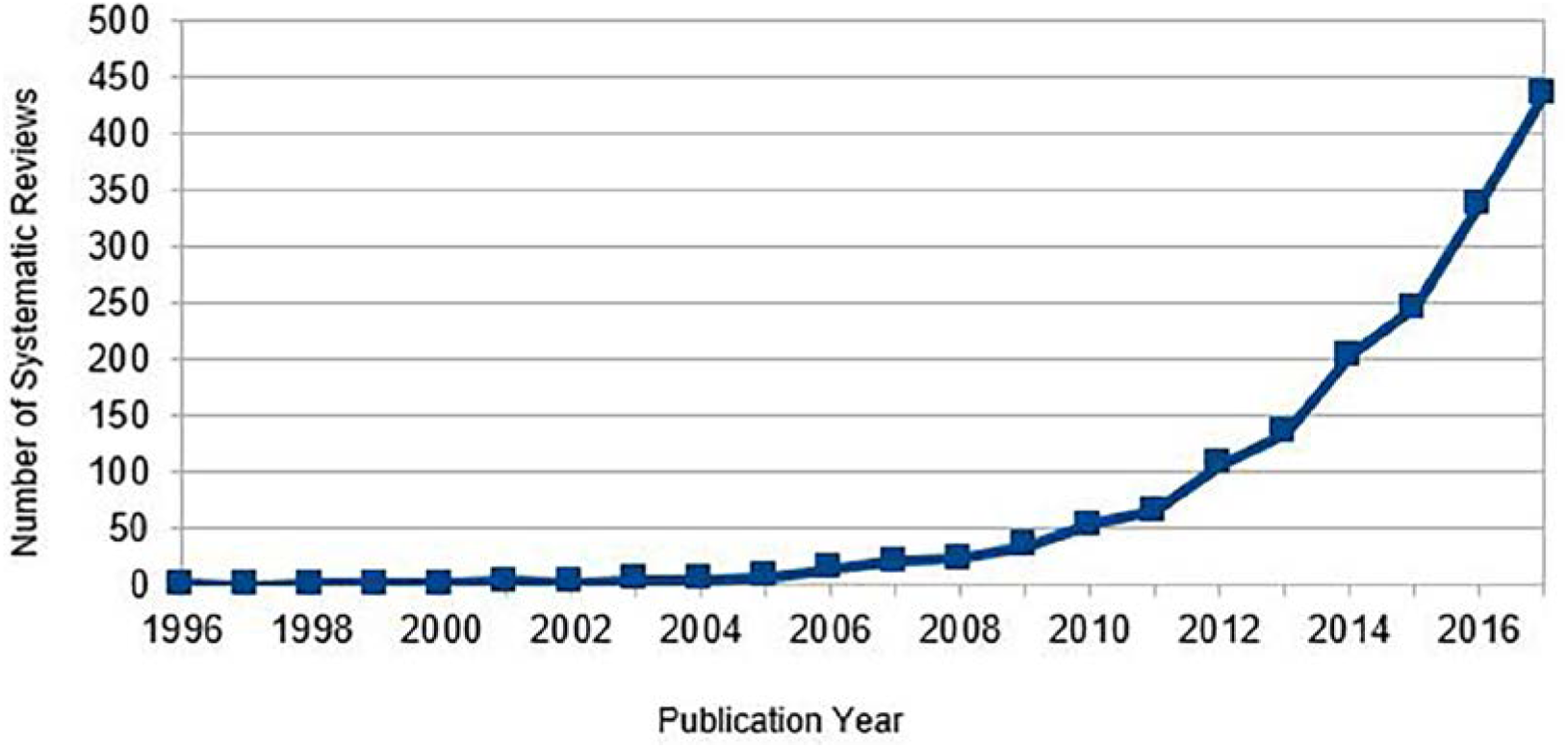
Number of librarian co-authored SRs published over time.

### Characteristics of articles citing librarian co-authored SRs

The total number of records for articles citing librarian co-authored SRs retrieved in the WOS Core Collection was 42,831 with self-citations (42,641 without self-citations), of which 41,610 with self-citations were available for export based on our institution-specific subscription. Therefore, the collective number of articles citing the 1,711 librarian co-authored SRs potentially available for analysis was 41,610. Of these records, 37,108 were found in PubMed, of which 28,868 were indexed in MEDLINE and available for publication type analysis. These citing articles were published between 1997 and 2018.

The 28,868 MEDLINE-indexed articles citing librarian co-authored SRs represent an array of publication types (Table 2). An expanded version of Table 2 showing the publication types included in “Other” is available in the Appendix. Note that the publication type descriptors may appear in more than one Publication Characteristics hierarchy category (Publication Components, Publication Formats, Study Characteristics, and Support of Research), and a single MEDLINE record may be assigned more than one publication type. The top 5 leading Publication Formats found for the citing articles to the SRs include: Journal Article, Review, Comment, Editorial, Letter, Practice Guideline, and Consensus Development Conference. The breakdown of the top 5 Study Characteristics include Randomized Controlled Trial, Meta-analysis, Comparative Study, Multi-center study, and Editorial.

**Table 2.**
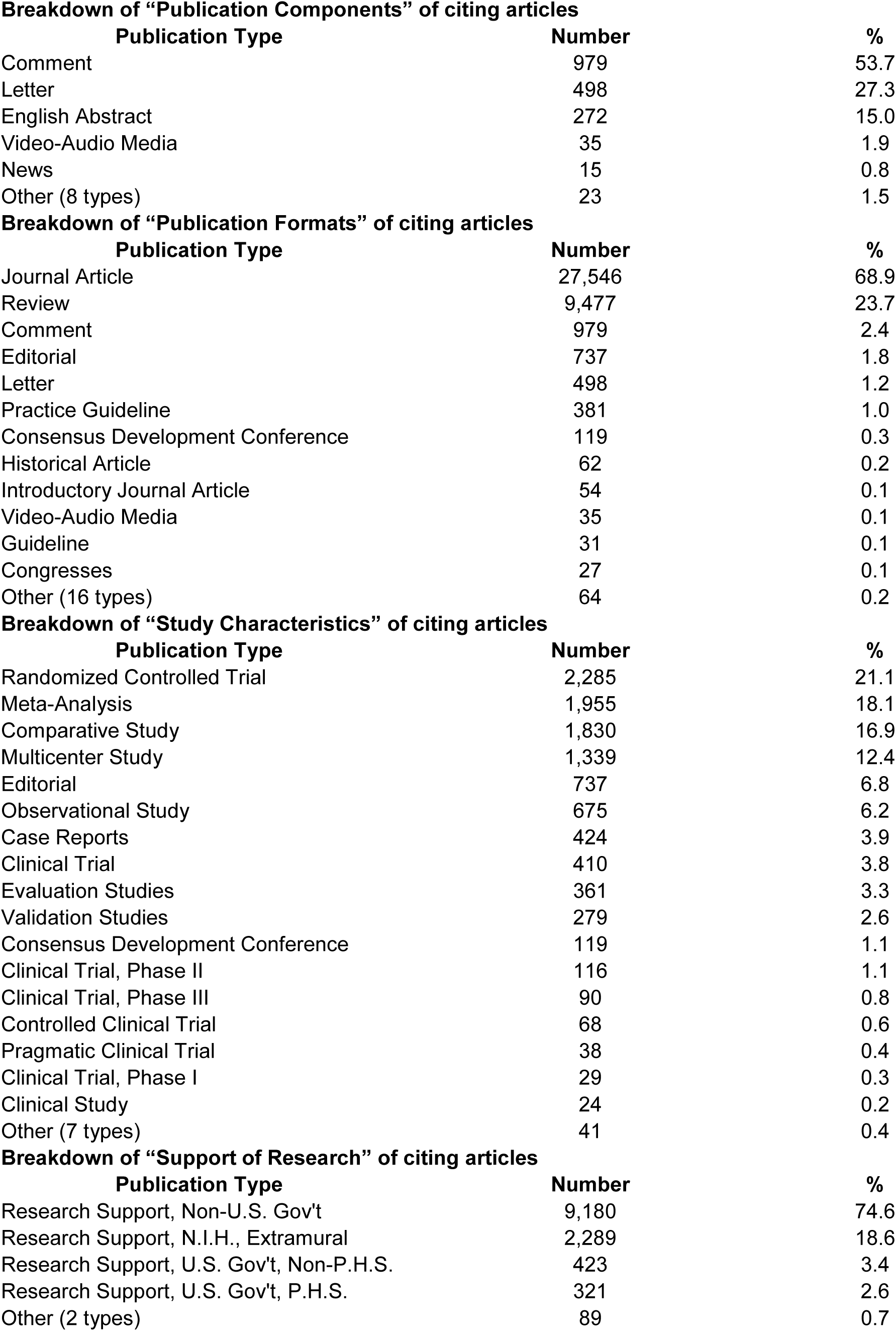
Publication characteristics of MEDLINE-indexed articles citing librarian co-authored SRs.

**Breakdown of “Publication Components” of citing articles**
| **Publication Type** | **Number** | **%** |
| --- | --- | --- |
| Comment | 979 | 53.7 |
| Letter | 498 | 27.3 |
| English Abstract | 272 | 15.0 |
| Video-Audio Media | 35 | 1.9 |
| News | 15 | 0.8 |
| Other (8 types) | 23 | 1.5 |

**Breakdown of “Publication Formats” of citing articles**
| **Publication Type** | **Number** | **%** |
| --- | --- | --- |
| Journal Article | 27,546 | 68.9 |
| Review | 9,477 | 23.7 |
| Comment | 979 | 2.4 |
| Editorial | 737 | 1.8 |
| Letter | 498 | 1.2 |
| Practice Guideline | 381 | 1.0 |
| Consensus Development Conference | 119 | 0.3 |
| Historical Article | 62 | 0.2 |
| Introductory Journal Article | 54 | 0.1 |
| Video-Audio Media | 35 | 0.1 |
| Guideline | 31 | 0.1 |
| Congresses | 27 | 0.1 |
| Other (16 types) | 64 | 0.2 |

**Breakdown of “Study Characteristics” of citing articles**
| **Publication Type** | **Number** | **%** |
| --- | --- | --- |
| Randomized Controlled Trial | 2,285 | 21.1 |
| Meta-Analysis | 1,955 | 18.1 |
| Comparative Study | 1,830 | 16.9 |
| Multicenter Study | 1,339 | 12.4 |
| Editorial | 737 | 6.8 |
| Observational Study | 675 | 6.2 |
| Case Reports | 424 | 3.9 |
| Clinical Trial | 410 | 3.8 |
| Evaluation Studies | 361 | 3.3 |
| Validation Studies | 279 | 2.6 |
| Consensus Development Conference | 119 | 1.1 |
| Clinical Trial, Phase II | 116 | 1.1 |
| Clinical Trial, Phase III | 90 | 0.8 |
| Controlled Clinical Trial | 68 | 0.6 |
| Pragmatic Clinical Trial | 38 | 0.4 |
| Clinical Trial, Phase I | 29 | 0.3 |
| Clinical Study | 24 | 0.2 |
| Other (7 types) | 41 | 0.4 |

| **Publication Type** | **Number** | **%** |
| --- | --- | --- |
| Research Support, Non-U.S. Gov't | 9,180 | 74.6 |
| Research Support, N.I.H., Extramural | 2,289 | 18.6 |
| Research Support, U.S. Gov't, Non-P.H.S. | 423 | 3.4 |
| Research Support, U.S. Gov't, P.H.S. | 321 | 2.6 |
| Other (2 types) | 89 | 0.7 |

## DISCUSSION

The rise in the number of SRs co-authored by librarians over time is consistent with professional practice and the evolving nature of services provided by health sciences libraries over the last decade [21]. Our findings shed light on the extent and nature of the scholarly contributions of librarian collaborators on SRs and constitute a benchmark for future research.

Our analysis revealed that the rate of librarian co-authored SRs appeared to accelerate around 2011. This coincides with the year the IOM SR standards were released, which includes a standard to work with a librarian (Standard 3.1) [1]. The publication of these standards may have positively impacted the number of SR service requests received by librarians, leading to an increased publication rate of librarian co-authored SRs. A related reason for this increased publication rate could be that librarians began advocating more aggressively for authorship. Supporting resources such as the IOM standards, studies showing that librarian involvement improves the quality of SRs [6-8], and librarian authored publications in leading health sciences journals advocating for librarians as members of SR teams [22] may have all played a role in facilitating librarian co-authorship. In tandem with librarian advocacy, increased researcher adherence to authorship guidelines [23] may also have contributed to the increased number of librarian co-authored SRs.

Co-authorship demonstrates value in expertise and the resulting collaboration, and partnerships with research groups. It is encouraging to see that librarians are increasingly being offered co-authorship for their contributions. This trend is particularly encouraging in light of other studies showing that librarians involved in SRs are often not included as authors, or only included in article acknowledgments, despite being deeply involved and making significant contributions to the SRs [8].

Our density visualization map shows that many librarian co-authored SRs addressed diagnostic test accuracy, as well as interventions, which are traditionally the most common clinical foci for SRs. This is consistent with the history of the Cochrane Collaboration, which originally developed only SRs on interventions but which began to develop SRs of diagnostic test accuracy in 2007 [24]. In addition, this map indicates that cancer and cardiovascular disease are the two disease categories most highly represented by librarian co-authored SRs. This is consistent with data from *National Vital Statistics Reports* and *Statistics Canada* indicating that cardiovascular-and cancer-related diseases are the top two leading causes of death in the United States and Canada [25, 26].

We found that librarian co-authored SRs were cited an average of 26.4 times, consistent with previous studies showing high citation rates for SRs and meta-analyses [27, 28]. This is also consistent with the mean citation rate of 26.5 over 4 years reported by Royle et al. in their bibliometric analysis of SRs [12]. We found that 10.9% of librarian co-authored SRs were uncited, which included the WOS document types “article,” “review,” “meeting abstract,” and “proceedings paper.” This percentage is higher than those reported by Royle et al. (1.6%) [12] and Van Noorden (4%) [29] and is likely a result of including meeting abstracts in our study. In our sample, the uncited rate for articles and reviews, excluding meeting abstracts and proceedings papers, was 3.8%, which is closer to the aforementioned studies’ uncited literature rates [12, 29]. There are many reasons for articles not being cited, not all of them negative [29]. These include the use of a single source (e.g., Web of Science Core Collection) for citation data and insufficient time for the dissemination of research. Of the 10.9% of uncited SRs, 41.7% were from 2017, indicating that insufficient time may have passed for these papers to receive citations.

Librarian co-authored SRs had an average of 7 authors, similar to that reported by Royle et al. for their top 50 most cited papers (6 authors) [12]. The top countries in which librarian co-authored SRs were most often published were the United States, Canada, Netherlands, England, China, and Australia. These results match the leading countries publishing SRs identified by Fontelo et al. in their PubMed analysis of evidence-based publication trends across countries which were, in rank order, the United States, United Kingdom, China, Australia, Canada, and Netherlands [30].

Not surprisingly, librarian co-authored SRs were most often published in the *Cochrane Database of Systematic Reviews*. The Cochrane Collaboration is a SR-producing entity, and Trial Search Coordinators/Information Specialists are routinely included as SR team members. One specialty journal, the *Journal of Clinical Endocrinology & Metabolism*, was among the top five sources of librarian co-authored SRs, which aligns with our density visualization map showing diabetes as a prominent research area. In our study, the leading journals publishing the top 10% most cited SRs included: *BMJ, Cochrane Database of Systematic Reviews, JAMA*, and the *Journal of Clinical Endocrinology & Metabolism*. The presence of the *Journal of Clinical Endocrinology & Metabolism* in this list is consistent with the Overlay Visualization Map (Figure 3), which shows “diabetes” as one of the higher citation impact areas among the research areas represented on the map.

In addition to analyzing librarian co-authored SRs themselves, we also conducted content analysis of a large number (>28k) of citing articles to those reviews. Our aim was to provide a more narrative view of impact by demonstrating the nature of librarian co-authored SRs’ diffusion within the scholarly literature. In the literature, The Becker Model provides a framework for research impact assessment that goes beyond traditional citation analysis to quantitatively demonstrate impact as a complement to citation impact [10]. Elements of the Becker model for demonstrating impact include, for example, evidence of knowledge transfer by way of a research study being cited in a meta-analysis, consensus development conference, patent, etc. and evidence of clinical implementation (defined as “the application or adoption of research outputs in clinical practice” [10]) such as a research study cited in guidelines, continuing education materials, or private and public health care benefit plans, etc.

In our study, 412 citing articles were characterized as Practice Guidelines and Guidelines. This demonstrates that librarian co-authored SRs were disseminated into guidelines with the potential for clinical implementation. While we did not examine the nature of how the librarian co-authored SRs were used in these citing sources, it is possible they were cited as part of the body of evidence to support a recommendation in the guidelines thus serving as surrogate for clinical implementation, as defined by the Becker Model. The diffusion of the SRs into the various study types suggests that the librarian co-authored SRs in this study were used to influence future research studies. Our study found that citing sources to librarian co-authored SRs were characterized as 2,285 Randomized Controlled Trails, followed by 1,955 Meta-analyses, 1,830 Comparative Studies, and 1,339 Multicenter Studies, to name a few of the top “Study Characteristics” Publication Types. These examples of our results along with the detailed list in Table 2, additionally demonstrate the impact of librarian co-authored SRs in a more descriptive manner which complements the citation analysis we conducted.

Librarian SR services provide opportunity for collaboration, deeper engagement, and education. Despite many challenges related to librarians providing SR support, various strategies can be employed by library administrators to incorporate SR services and allay concerns. The results of the present study could be useful for health sciences library departments and for institutions to demonstrate the value of librarian services for SRs by highlighting librarian contributions to research and practice in other disciplines. Librarians on tenure/tenure-equivalent tracks may face challenges in convincing their tenure and promotion committees that non-LIS SR scholarship is equivalent to LIS scholarship. However, our study provides evidence of the impact of librarian scholarship on the knowledgebase of non-LIS disciplines, which requires expertise that is uniquely within the skill set of librarians. Thus, our findings may be useful in expanding the definition of scholarship for academic librarians.

### Limitations

There are many challenges associated with conducting bibliometric analyses, including indexing, sources used, and retrieval of the information discovered, all contributing to the difficulty of providing a comprehensive analysis. A limitation of this study is that we may have missed librarian co-authored SRs due to our use of WOS as a single source and our reliance on author indexing within this source. Solely using the term “library” and its variants in our search may have also limited our results, as some libraries do not use the term “library” in their name, and some librarians work in non-library contexts or have primary or co-appointments in other institutional departments. Another limitation is that we were not able to analyze the entire corpus of citing articles in WOS Core Collection because the records available for export were dependent on our institutional subscription. Additionally, citing articles did not comprehensively encompass grey literature, which is not well indexed in traditional bibliographic databases. Finally, while we identified the publication types of MEDLINE-indexed citing articles, we do not know how they were used in the publications; this would require more in-depth analysis of the full text of the literature, which was not feasible with the large scale of citing articles we analyzed.

### Further Research

Our results could inspire several directions for further research. For instance, it would be valuable to examine SRs for different types of outcomes other than scholarship. It could be also be valuable to do more in depth content analysis to investigate how citing literature use the cited librarian co-authored SRs.

## CONCLUSION

Our characterization of the scholarly outputs and impact of librarian co-authored SRs emphasizes the value of the unique SR expertise of librarians. SRs are research that can have significant impact in many areas such as policy, patient care, and further research [31]. That librarians are increasingly providing SR services, despite the challenges involved, strengthens the vital role that librarians can play in research overall and could help expand the definition of scholarship within the LIS profession. This study may serve as a benchmark for administrative decision making about library SR services and for future research in this area.

## Data Availability

Data will be made available in a open access repository. Submission record will be updated with data identifiers when the data becomes available.

## Acknowledgements

The authors would like to thank the various colleagues who provided helpful comments in review of this manuscript. The authors are also grateful for the invaluable assistance provided by Ludo Waltman, Yvonne Mery, and Carol Howe.

## APPENDIX

Table 2 (expanded version) Publication characteristics of the MEDLINE citing articles to the 1,711 SRs.

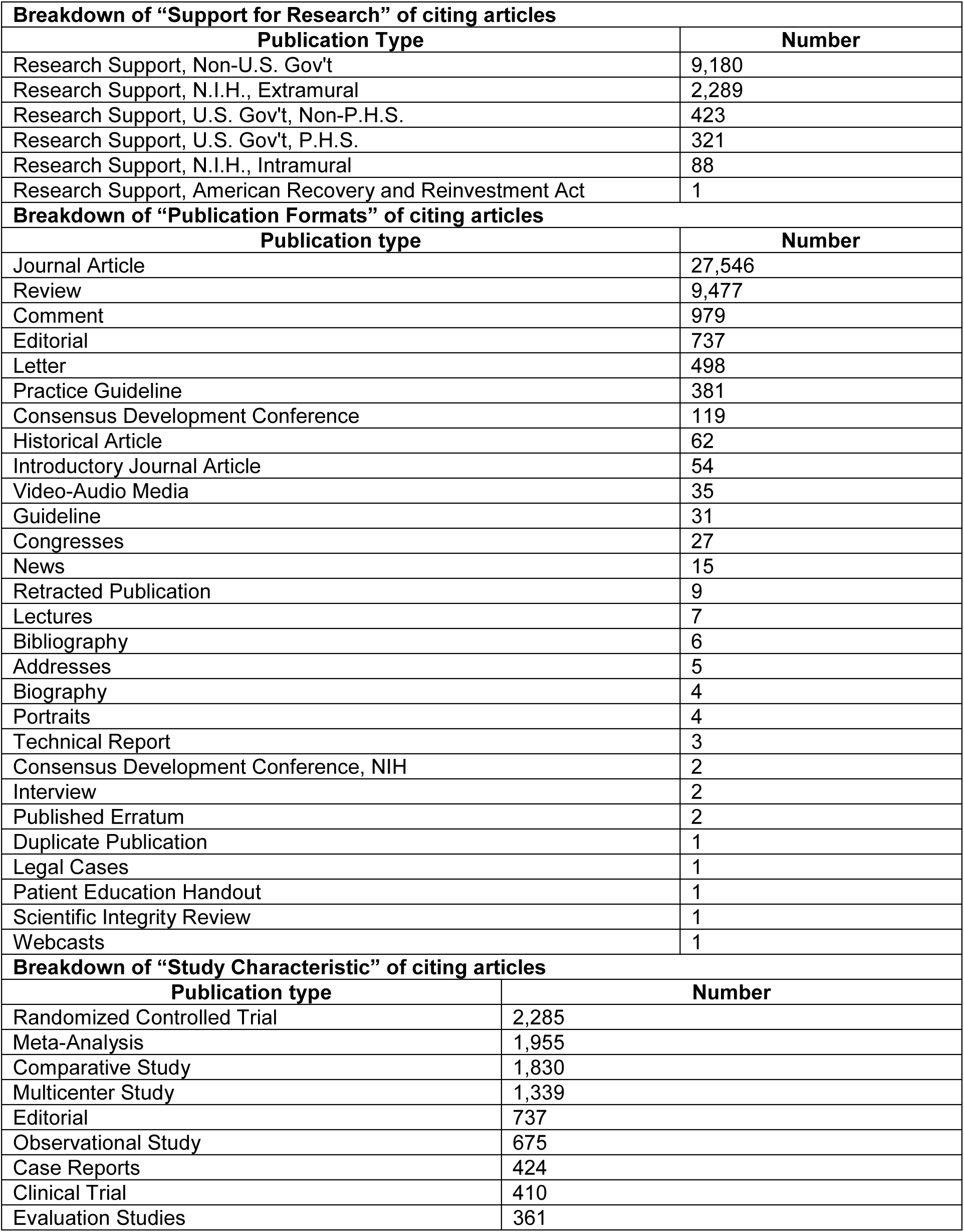
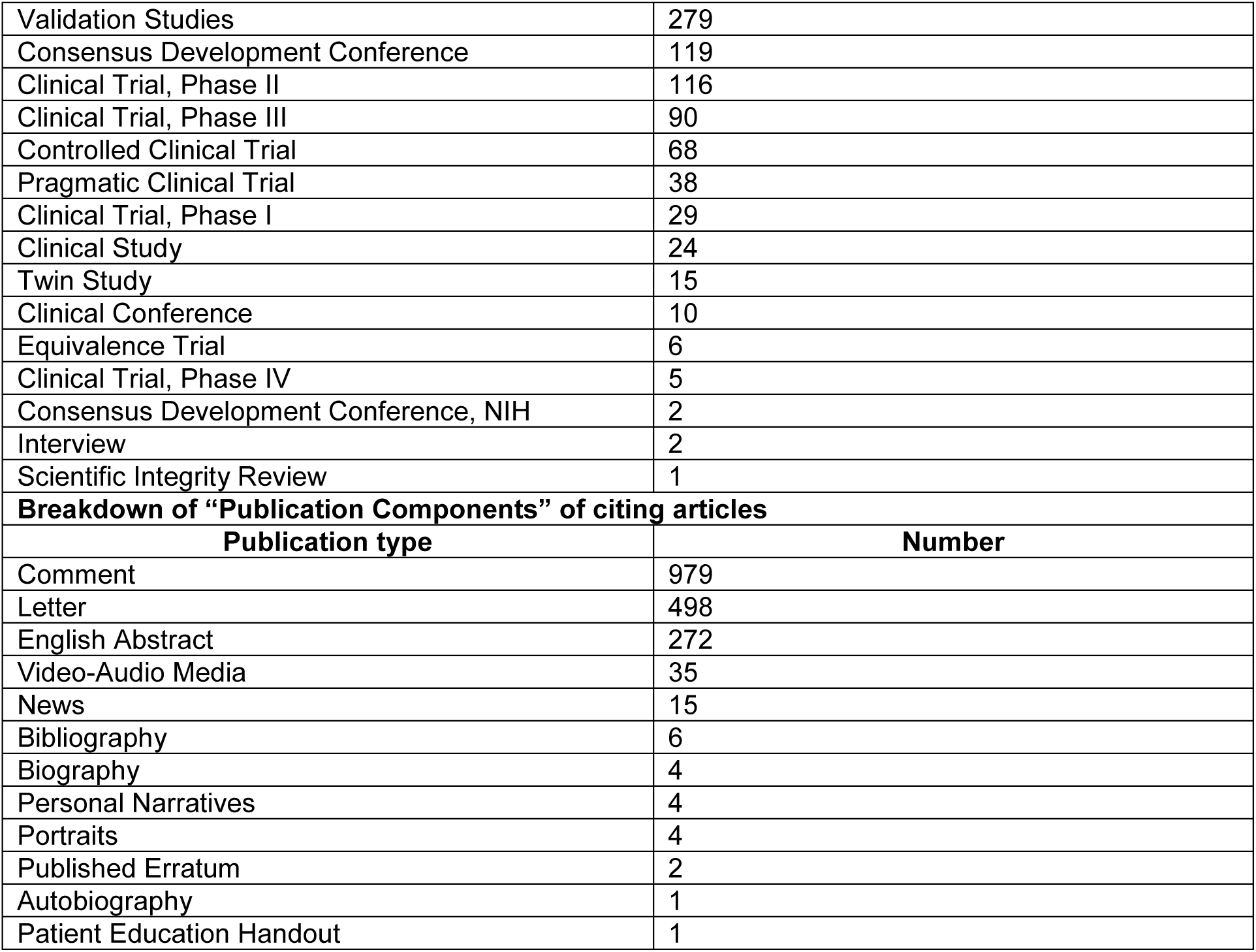

